# The Prognostic Role of *IRS2* in Breast Cancer: Interactions with Glucose Metabolism and Menopausal Status

**DOI:** 10.1101/2025.07.14.25331512

**Authors:** Katie Liu, Aditya Shah, Gautham Ramshankar, Rachel Perry

**Author notes:** Corresponding author: Rachel J. Perry, Ph.D., Correspondence to; 333 Cedar St. SHM BE36-B, New Haven, CT 06520 U.S.A.

## Abstract

The Insulin Receptor Substrate 2 (*IRS2*) gene encodes an adapter protein that transmits insulin signals to regulate various cell functions, including metabolism, growth, and survival. Dysregulation of IRS2 activity is often associated with obesity and type 2 diabetes. Both conditions are strongly associated with a higher breast cancer risk. IRS2 is predominantly regulated in various tissues at the level of expression; however, the association between tumor *IRS2* expression and certain clinical factors, such as diabetes status and menopausal status, and their effects on breast cancer prognosis are not fully understood. To address this gap, we utilized five datasets of bulk RNA-seq data of patients with breast cancer from the Xena Functional Genomics Explorer, and primarily analyzed the TCGA Breast Cancer Cohort, which includes data from 1097 patients with primary breast tumors. We complemented these transcriptomic analyses by using 3′-Deoxy-3′-18F-Fluorothymidine positron emission tomography-computed tomography (^18^F-FLT PET-CT) scans from 23 patients that were publicly available from the Cancer Imaging Archive (TCIA) to correlate this marker of tumor cell proliferation with body mass index (BMI) and glucose levels (as a proxy for diabetes), and further divided data by menopausal status. Through deidentified, publicly available data, the study explores the effects of *IRS2* expression on breast cancer in different cohorts, as well as the role of menopausal status in this dynamic and the context of obesity and diabetes.

## Introduction

With around 2.26 million cases reported in 2020, breast cancer is the most commonly diagnosed cancer in the world [1,2]. Similar to many other diseases, breast cancer risk is shaped by both genetic and environmental factors. Among modifiable risk factors for breast cancer, diabetes and obesity stand out due to their increasing global prevalence and their known, but not completely defined mechanistic relationship with breast cancer [3,4]. These factors not only affect cancer risk but also influence outcomes, making it pressing to understand the metabolic and molecular components that contribute to this tangled web linking obesity, diabetes, and breast cancer, in which insulin signaling is a key participant [4].

The gene Insulin Receptor Substrate 2 (*IRS2*) gene encodes insulin receptor substrate 2 (IRS2), an adapter protein that transmits insulin signals to regulate glucose metabolism and cell growth [5]. A dysregulation of IRS2 activity has a strong association with obesity and type 2 diabetes by contributing to systemic insulin resistance [6,7]. Mice lacking *IRS2* failed to produce β-cell hyperplasia or increase insulin secretion to compensate for the absence of insulin signaling in metabolically active tissues, and they developed type 2 diabetes [8]. In contrast, the absence of *IRS1*, another isoform of the insulin receptor substrate, did not affect glucose tolerance due to compensatory β-cell hyperplasia, consistent with the idea that a failure of β-cell hyperplasia in response to insulin resistance may be the primary defect causing the development of type 2 diabetes [8]. Additionally, the absence of *IRS3* and *IRS4* both induced minimal disruptions to glucose metabolism, suggesting that IRS2 is a primary substrate of insulin signaling and, hence, a vital factor in insulin resistance and type 2 diabetes, and interesting to study in the context of breast cancer [8]. Higher levels of *IRS2* expression are observed in cancer cells compared to normal tissues, suggesting a role in tumorigenesis through the activation of the IRS-PI3K-AKT and MAPK pathways [9,10].

As diabetes and cancer both share overlapping metabolic underpinnings marked by alterations in glucose metabolism, it is not surprising to see that insulin signaling is at the center of some of the metabolic shifts implicated in cancer [3]. However, menopausal status contributes to a more complex link between these two factors. As women go from a premenopausal to a postmenopausal state, they experience a decrease in estradiol and an increase in estrone, leading to abdominal fat accumulation and obesity, which is expected to impair insulin sensitivity [11,12]. Higher estrogen levels directly increase estrogen receptor-positive breast cancer risk, and postmenopausal women with obesity have higher estrogen levels compared to their non-obese counterparts. In women who are obese, adipose tissues can increase estrone production due to the activity of aromatase, an enzyme that converts androgens into estrogens [13,14]. However, obesity causes a negative feedback in the hypothalamic-pituitary axis for premenopausal women, resulting in reduced gonadotropin secretion [15]. Anovulation can occur in premenopausal women with obesity, thereby reducing levels of estrogens as compared to normally cycling, normal weight women, and this reduced exposure to gonadotropins could support the inverse relationship between obesity and breast cancer risk in premenopausal women, not experiencing the same change in insulin sensitivity as postmenopausal women [16]. Due to this, we anticipate that menopausal status and glucose metabolism would interact to affect breast cancer risk, and that menopausal status would moderate this interaction.

Positron emission tomography/computed tomography (PET/CT) is an imaging method that combines PET and CT scans to provide a comprehensive series of images of the body’s anatomical structure and metabolic activity [17]. Due to this, it is commonly used in oncology for various types of cancer [17–20]. It has become a very reliable tool for staging metastatic breast cancer [20]. This method measures uptake of [^18^F]-3’-fluoro-3’-deoxythymidine (^18^F-FLT, a radiotracer) to demonstrate the amount of thymidine accumulation in both primary tumor tissues and metastatic lesions. Higher ^18^F-FLT uptake indicates greater thymidine accumulation for DNA synthesis and, consequently, more proliferation [20,21]. The uptake of the radiotracer is measured as the standard uptake value (SUV), the numerical amount of radiotracer absorbed by tissues, making it a reliable quantitative metric for cell proliferation. Additionally, blood glucose levels are a marker of a patient’s status on the continuum between normal glucose metabolism and diabetes [22], and were incorporated into this study to reflect a common readout of metabolic dysfunction.

To illustrate the association between IRS2 and breast cancer outcomes using a larger sample size than the TCIA image analysis, the RNA-seq data in the UCSC Xena Functional Genomics Explorer were utilized. This tool allowed for the visualization of how *IRS2* expression affects breast cancer prognosis in premenopausal and postmenopausal patients. Our analyses highlight novel insights into the relationship between clinical variables (menopausal status, glucose metabolism), cell proliferation, tumor *IRS2* expression, and survival rates of breast cancer patients, which may support interventional studies focusing on these variables in patients with breast cancer.

## Methods

### *IRS2* gene expression analysis

The TCGA Breast Cancer (BRCA) dataset was input into the UCSC Xena Functional Genomics Browser’s Visualization tab to investigate breast cancer prognosis. (Dataset Link: https://xenabrowser.net/datapages/?dataset=TCGA.BRCA.sampleMap%2FBRCA_clinicalMatrix&host=https%3A%2F%2Ftcga.xenahubs.net&removeHub=https%3A%2F%2Fxena.treehouse.gi.ucsc.edu%3A443). There were 1247 total patients. Since Column A is automatically formatted by UCSC for all their datasets, we did not omit any data. Column B was selected as a genomic data type, *IRS2* was selected as the gene, and gene expression was chosen as the dataset. Column C was selected as a phenotypic data type, and menopause status was chosen as the phenotype. All null and duplicate samples were removed, resulting in 1097 samples of the original 1247 remaining for analysis. A Kaplan-Meier (KM) plot was created for Column B to visualize IRS2 expression and its impact on prognosis. Next, we separated the samples into low- and high-expression groups. After removing the 34 samples with indeterminate menopausal statuses, Xena determined 8.867 FPKM to be the median for IRS2 expression, serving as the dividing point between high and low expression groups. There were 485 patients in the low-expression group (<8.867 FPKM) and 486 patients in the high-expression group (>=8.867 FPKM). A KM plot was created for both groups, using Column C to reveal menopausal status (premenopausal, postmenopausal, and perimenopausal) and its effects on prognosis in each expression group.

Additional breast cancer patients’ tumor gene expression data was accessed through the following data sets: the Breast Cancer (Caldas 2007) [23] (https://xenabrowser.net/datapages/?cohort=Breast%20Cancer%20(Caldas%202007)&removeHub=http%3A%2F%2F127.0.0.1%3A7222), the GDC TCGA Breast Cancer (BRCA) (https://xenabrowser.net/datapages/?cohort=GDC%20TCGA%20Breast%20Cancer%20(BRCA)&removeHub=http%3A%2F%2F127.0.0.1%3A7222), the Breast Cancer (Miller 2005) [24] (https://xenabrowser.net/datapages/?cohort=Breast%20Cancer%20(Miller%202005)&removeHub=http%3A%2F%2F127.0.0.1%3A7222), and the Node-negative breast cancer (Desmedt 2007) [25] (https://xenabrowser.net/datapages/?cohort=Node-negative%20breast%20cancer%20(Desmedt%202007)&removeHub=http%3A%2F%2F127.0.0.1%3A7222). We examined breast cancer datasets that included tumor *IRS2* gene expression and survival data to create KM plots on the Xena platform. None of the datasets had information on menopausal status. For each dataset, *IRS2* was added to Column B as a genomic data type, null and duplicate samples were removed, and a KM plot was generated. Overall Survival was set as the dependent variable for all KM plots unless otherwise specified.

### ^18^F-FLT PET-CT image analysis

Deidentified PET-CT images were acquired on The Cancer Imaging Archive (TCIA) from the ACRIN 6688 clinical trial. The “ACRIN-FLT-Breast (ACRIN 6688)” dataset can be found here: https://www.cancerimagingarchive.net/collection/acrin-flt-breast/). The all-adult participant group provided written consent for their data to be used in public repositories when de-identified, making separate ethical approval for this dataset and other datasets used in this manuscript unnecessary. The University of Arkansas for Medical Science (UAMS) Institutional Review Board (IRB #205568) approved data sharing via TCIA, and the patients gave informed consent for their data to be shared with TCIA. We do not have access to the details of the consent process. Separate IRB approval was not necessary because our use of this de-identified data was already approved under these IRB conditions.

All scans of patients with a menopausal status, diabetes status, height, weight, and five clear CT slices (slices that allowed for the primary breast tumor to be identified and for its ^18^FLT-SUV values to be generated) were included for analysis. Data from nine premenopausal and fourteen postmenopausal patients met these conditions, and all were analyzed. If patients had multiple scans, only the earliest scan was analyzed to minimize the effect of neoadjuvant chemotherapy on the tumor. All applicable scans were uploaded to Fiji ImageJ, and the PET-CT Viewer plugin was used to complete the analysis. Once a slice that included an identifiable primary breast tumor was found, fixed volume spheres were drawn around the interior regions of interest (ROIs) on that CT slice using the Brown Fat Volume tool. The mean thymidine uptake in the tumor tissue was measured from these fixed volume spheres, with only two criteria selected: SUV parameters (2-15) and the “any” voxel criteria. Consequently, the lean body mass-corrected standardized uptake values (SUV) were revealed. This was repeated until data from five slices were collected for each patient. The primary objective was to determine the SUV (g/mL) of tumors correlated with diabetes status (glucose levels in mg/dL) and obesity status (BMI values in kg/m^2^).

GraphPad Prism was used to analyze data. For all datasets, a Shapiro-Wilk normality test was performed. For the datasets where both variables were continuous, not all analyses were found to be normally distributed. As a result, two separate significance tests were conducted. If the analysis was found to be normally distributed, a two-sided Pearson correlation test was performed to determine significance. If the analysis was found to be not normally distributed, a two-sided Spearman correlation test was performed to assess significance. For the datasets with one categorical and one continuous variable, all datasets were found to be normally distributed. As a result, a Welch’s t-test was performed to compare groups.

## Results

### *IRS2* expression and survival probability

By comparing the patients in the high *IRS2* expression group to the low IRS2 expression group in the TCGA Breast Cancer (BRCA) dataset, we come across the following findings: from 0 days to 3500 days, patients in the high *IRS2* expression group had a higher survival rate compared to those in the low *IRS2* expression group; from 3500 days to 6250 days, the patients in the low *IRS2* expression group had a higher survival rate; and from 6250 days to the end of the study, patients in the high expression group had a higher rate of survival (Fig 1A). In like manner, from 0 days to 3500 days, patients in the high *IRS2* expression group had a higher disease-specific survival rate than those in the low *IRS2* expression group, except 2750 days to 3000 days; from 3500 days to 4500 days, patients in the low *IRS2* expression group had a higher disease-specific survival rate. However, from 4500 to 6500 days, patients in the high *IRS2* expression group had a higher disease-specific survival rate; from 6500 days to the end of the study, patients in the low expression group had a higher disease-specific survival rate (Fig. 1B). The tendency for *IRS2* to exhibit differing effects on survival based on days after diagnosis suggests the possibility of a differing impact of *IRS2* based on cancer stage. The datasets we used did not include a staging timeline, but future studies could be designed to assess this. While we recognize that the long follow-up period impedes our ability to attribute mortality to breast cancer fully, *IRS2* holds promise as a valuable prognostic factor, potentially requiring further segmentation by tumor stage, treatment, and/or other clinical characteristics.

**Fig 1.**
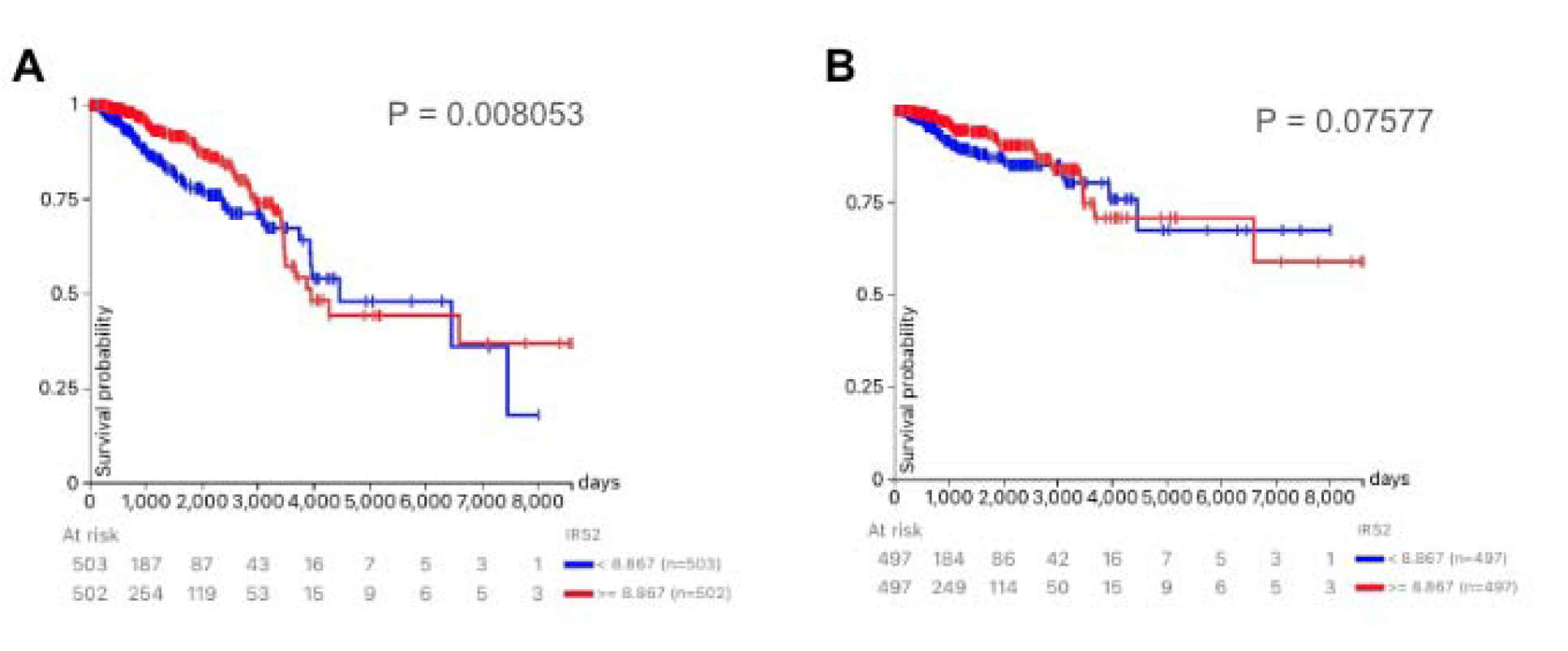
*IRS2* expression affects survival probability of breast cancer patients. Survival of patients in the TCGA BRCA database with high expression of IRS2 (>= 8.867 FPKM) vs low expression of *IRS2* (<8.867 FPKM) in the TCGA Breast Cancer (BRCA) dataset, in regards to (A) overall survival probability and (B) disease-specific survival probability.

In contrast to the data from the TCGA Breast Cancer dataset, in the “Breast Cancer (Caldas 2007)” gene expression dataset, the patients in the high *IRS2* expression group (>=0.02978 log2) had very similar survival rates to the patients in the low *IRS2* expression group (<0.02978 log2) (Fig 2A). In the “GDC TCGA Breast Cancer (BRCA)” gene expression dataset, from 0 days to 3500 days, patients in the high *IRS2* expression group (>=2.102 FPKM) had a higher survival rate than the low *IRS2* expression group (<2.102 FPKM); from 3500 days to 6500 days, the patients in the low *IRS2* expression group had a higher survival rate than the high *IRS2* expression group; and from 6500 days to the end of the study, patients in the high *IRS2* expression group had a higher survival rate than the low *IRS2* expression group (Fig 2B). In the “Breast Cancer (Miller 2005)” gene expression dataset, patients in the high *IRS2* expression group (>=−0.05655 log2) had a higher survival rate compared to patients in the low *IRS2* expression group (<−0.05655 log2) for the entirety of the study (Fig. 2C). In the “Node-negative breast cancer (Desmedt 2007)” gene expression dataset, from 0 days to 2000 days, patients in the low IRS2 expression group (<0.07265 log2) had a higher survival rate than patients in the high *IRS2* expression group (>=0.07265 log2). From 2000 days to the end of the study, patients in the high *IRS2* expression group had a higher survival rate (Fig. 2D). Overall, the datasets suggest that high *IRS2* expression is associated with similar, if not worse, prognosis for breast cancer patients compared to low *IRS2* expression, with two datasets demonstrating lower survival rates for patients in the high *IRS2* expression group and two datasets showing similar survival rates for the two groups.

**Fig 2.**
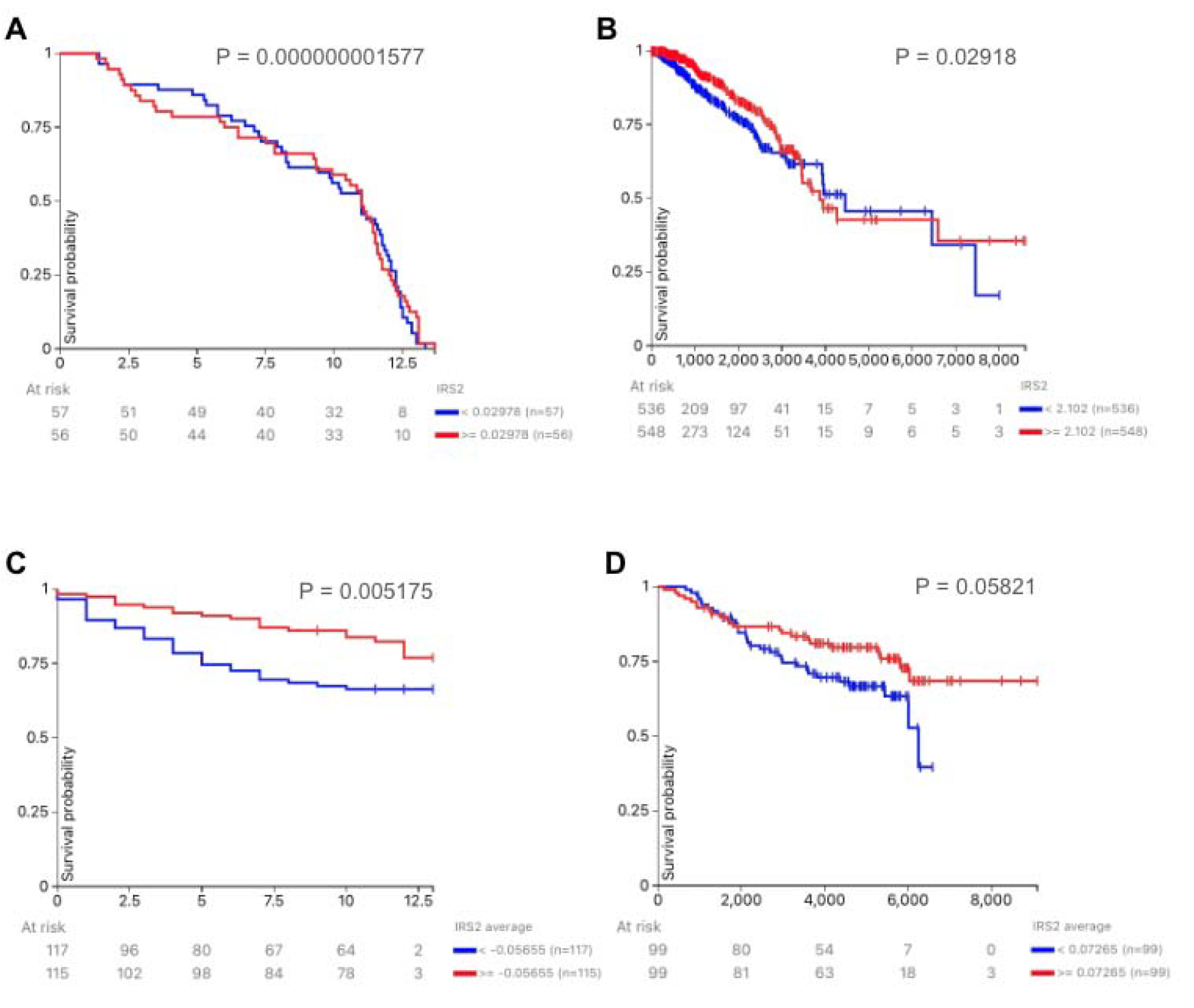
The impact of *IRS2* expression on overall survival probability in patients with breast cancer differs by dataset. Survival was examined in patients from the (A) “Breast Cancer (Caldas 2007)”, (B) “GDC TCGA Breast Cancer (BRCA)”, (C) “Breast Cancer (Miller 2005)”, and (D) “Node-negative breast cancer (Desmedt 2007)” by high vs low *IRS2* expression.

### Impact of menopausal status on survival probability in patients with breast tumors separated by high and low expressions of *IRS2*

Figure 3 illustrates the effect of menopausal status on overall survival in breast cancer patients with high and low *IRS2* expression. Premenopausal patients with high *IRS2* expression had a higher survival rate than postmenopausal patients in the high *IRS2* expression group. Perimenopausal patients in the high *IRS2* expression group had a survival rate of 100%, the highest of the three groups. Premenopausal patients in the low *IRS2* expression group had a marginally higher survival rate than postmenopausal patients in the low *IRS2* expression group.

**Fig 3.**
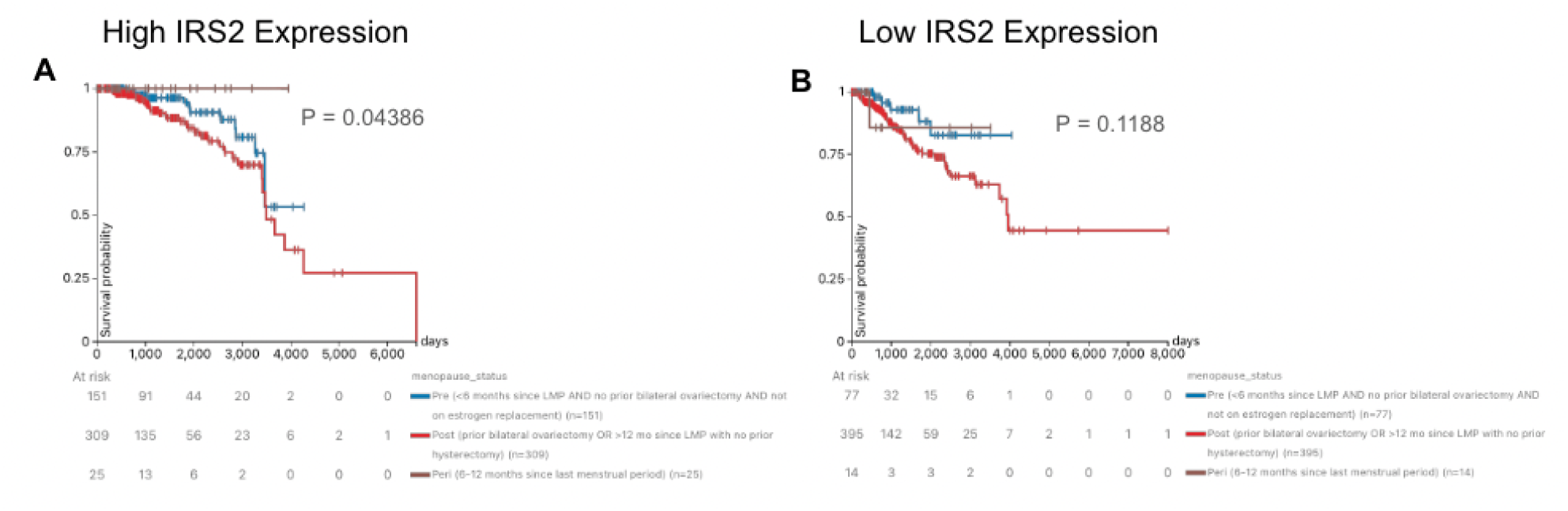
High and low *IRS2* expression in TCGA BRCA and prognosis by menopausal status. Prognosis for breast cancer patients with (A) high *IRS2* expression (>=8.867 FPKM) and (B) low *IRS2* expression (<=8.867 FPKM) in the TCGA BRCA dataset by menopausal status: premenopausal, postmenopausal, and perimenopausal.

Peri-menopausal patients in the low *IRS2* expression group tended to have the lowest survival rate. However, the low sample size of peri-menopausal patients limits our ability to draw conclusions for this group. Additionally, age is likely an important confounder: regardless of IRS2 expression, older patients with breast cancer, who are more likely to be postmenopausal, have increased all-cause mortality as compared to their younger counterparts, independent of their tumor’s characteristics [26].

### Correlations between blood glucose levels and tumor SUV mean separated by menopausal status

Prediabetic/diabetic patients exhibit a marginal tendency toward worse breast cancer prognosis compared to nondiabetic patients as assessed by tumor ^18^FLT-SUV_mean_ (Fig. 4A). Tumor SUV_mean_ is insignificantly positively correlated with blood glucose levels in premenopausal breast cancer patients (Fig. 4B). Similarly, tumor SUV_mean_ is insignificantly (approaching statistical significance) positively correlated with blood glucose levels in postmenopausal breast cancer patients Fig. 4C).

**Fig 4.**
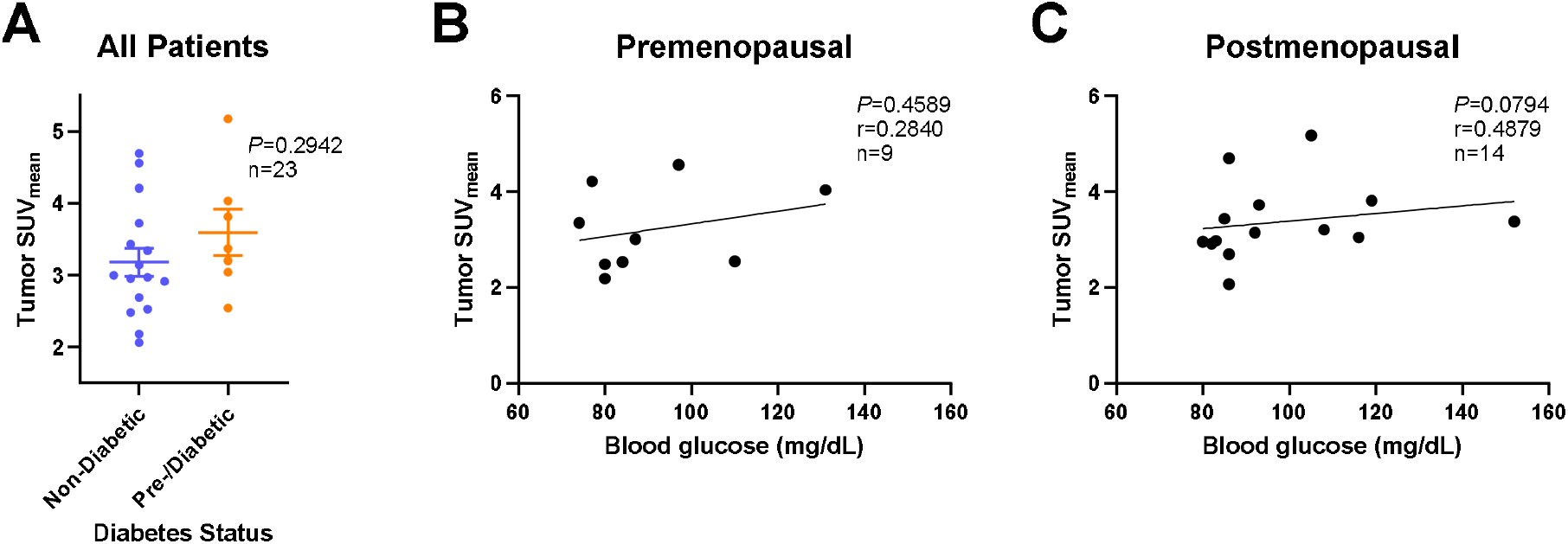
Lean body mass-corrected thymidine uptake separated by diabetes status. Correlation of diabetes status by ^18^FLT-SUV_mean_ in breast cancer patients. A Shapiro-Wilk normality test was performed. Because datasets were normally distributed, a Welch’s t-test was performed, and the p-value produced was used to determine statistical significance. Correlation of blood glucose levels with ^18^FLT-SUV_mean_ in breast cancer patients for (B) premenopausal patients and (C) postmenopausal patients. A Shapiro-Wilk normality test was performed. Based on normality, either a two-sided Pearson correlation or a two-sided Spearman correlation was performed, and the p-value produced from each test was used to determine statistical significance. SUV_mean_ - lean body mass-corrected thymidine uptake value.

## Discussion

IRS2 is known to play a key role in maintaining breast cancer stem cells, which can lead to primary or recurrent tumor growth even after drug treatment [27]. Previous studies have emphasized the involvement of IRS2 in insulin signaling, tumor cell proliferation, and breast cancer metastasis [5,10,27]. This study aims to expand the field’s understanding of the impact of insulin signaling via IRS2 on breast cancer prognosis when separated by menopausal status, obesity, and diabetes status, as there is limited information surrounding the interplay between these three factors in patients with breast cancer.

In our study, the impact of *IRS2* expression on survival outcomes is inconsistent, varying based on the dataset. When examining the TCGA BRCA dataset, the survival probability of patients alternated between high and low expression levels for both overall survival and disease-specific survival. Similarly, the “Breast Cancer (Caldas 2007)” dataset, the “GDC TCGA Breast Cancer (BRCA)” dataset, and the “Node-negative breast cancer (Desmedt 2007)” dataset all had the same fluctuation in survival trends between low and high *IRS2* expression. However, the “Breast Cancer (Miller 2005)” dataset had a better overall survival probability for the high *IRS2* expression group and the “Node-negative breast cancer (Desmedt 2007)” had a better overall survival probability for the high *IRS2* expression group for the majority of the period (past 2000 days), suggesting that high *IRS2* expression may have a slight advantage in terms of survival probabilities under certain conditions. The reason for this is unknown, but it could be attributed to external factors, such as hormone receptor status or recurrence [28], providing future researchers with possible factors that build upon gene expression and are worthy of investigation. Understanding this inconsistent trend reveals that there may be factors closely associated with *IRS2* expression, such as insulin signaling readouts, which largely occur at the level of enzyme phosphorylation, that have a greater impact on breast cancer prognosis than expression of *IRS2* itself. However, we were unable to investigate these other factors due to limited data availability.

Meaningful patterns for the impact of diabetes were confirmed when patients were sorted by diabetes status. Previous literature indicates that diabetes is a risk factor for breast cancer [3]. To explain, insulin resistance is a main cause for the development of diabetes [29]. To compensate for the lack of insulin function, the body produces more insulin, leading to hyperinsulinemia, a risk factor for breast cancer [30]. This may be further bolstered by the analysis in Fig. 4, which showed a higher ^18^FLT-SUV_mean_ for the prediabetic/diabetic subgroup compared to the normal subgroup. While the p-value indicated it as statistically insignificant, this could be attributed to a type II error due to the small sample size available. Tendencies for a meaningful impact of the impact of diabetes were also confirmed when patients were sorted by menopausal status. Previous literature has found that diabetes serves as a risk factor for cancer with no specification of menopausal status [3]. The data in Fig. 5 show that for both premenopausal and postmenopausal women, glucose levels had a positive correlation with tumor SUV_mean_, suggesting that diabetes worsens breast cancer prognosis regardless of menopausal status.

Past research has found that the absence of *IRS2* leads to insulin resistance, suggesting a positive relationship between the two [31]. Using RNA sequencing, we categorized patients into groups based on their *IRS2* expression status, and further segmented the data by menopausal status. We investigated survival probabilities for both high and low *IRS2* expression groups (Fig. 3). In the high expression group, premenopausal patients had overlapping survival probabilities with postmenopausal patients. In the low expression group, premenopausal patients had consistently higher survival probabilities compared to the postmenopausal patients. However, the low expression Kaplan-Meier plot had a statistically insignificant p-value. Further research can be conducted to determine if adjusting the expression threshold from 8.867 FPKM to a lower value yields statistically significant results within the low-expression group. Our results also support previous literature that has found that diabetes serves as a risk factor for breast cancer and for poorer survival outcomes in patients with breast cancer [3,32], although these results are often confounded by factors associated with both diabetes and cancer risk, such as advanced age [33,34]. This also bolsters the findings from the thymidine intake values as measured using the ^18^F-FLT PET-CT when results were separated into diabetes status, as well as when results were separated into menopausal status with blood glucose as an independent variable. This analysis indicated that higher blood glucose was marginally associated with higher ^18^F-FLT-SUV_mean_ values, supporting the gene expression results of perimenopausal patients experiencing lower survival rates when *IRS2* is not highly expressed. It is important to note that the premenopausal group and postmenopausal group both had varying survival outcomes, with neither maintaining a 100% survival rate, indicating it is not solely diabetes status that induced the difference in survival for perimenopausal patients and that their perimenopausal status played a role as well.

## Conclusion

Our analyses indicate that *IRS2* has a mixed effect on breast cancer prognosis depending on other clinical characteristics. These data highlight the importance of exploring other factors in relation to *IRS2*, such as insulin signaling, that may be more reliably associated with breast cancer prognosis. This conclusion is suggested by data from this study employing various clinical variables to correlate tumor proliferation and diabetes. Through this, we were able to identify that diabetes worsens breast cancer prognosis regardless of menopausal status, a specification that was not clear in previous literature. Future researchers should use larger sample sizes for correlation tests, which were not available for the current analyses, and perform interventional studies to ensure that findings are representative of larger populations. Additionally, interventions targeting *IRS2* expression and downstream signaling may be of interest in the future for patients with breast cancer.

## Data Availability

All data analyzed are available to anyone online at:
- https://xenabrowser.net/datapages/?dataset=TCGA.BRCA.sampleMap%2FBRCA_clinicalMatrix&host=https%3A%2F%2Ftcga.xenahubs.net&removeHub=https%3A%2F%2Fxena.treehouse.gi.ucsc.edu%3A443
- https://xenabrowser.net/datapages/?cohort=Breast%20Cancer%20(Caldas%202007)&removeHub=http%3A%2F%2F127.0.0.1%3A7222
- https://xenabrowser.net/datapages/?cohort=GDC%20TCGA%20Breast%20Cancer%20(BRCA)&removeHub=http%3A%2F%2F127.0.0.1%3A7222
- https://xenabrowser.net/datapages/?cohort=Breast%20Cancer%20(Miller%202005)&removeHub=http%3A%2F%2F127.0.0.1%3A7222
- https://xenabrowser.net/datapages/?cohort=Node-negative%20breast%20cancer%20(Desmedt%202007)&removeHub=http%3A%2F%2F127.0.0.1%3A7222
-https://www.cancerimagingarchive.net/collection/acrin-flt-breast/

